# Socioeconomic status and the risk for colonization or infection with priority bacterial pathogens: a global evidence map

**DOI:** 10.1101/2024.04.24.24306293

**Authors:** Sarah Blackmon, Esther Avendano, Nanguneri Nirmala, Courtney W. Chan, Rebecca A. Morin, Sweta Balaji, Lily McNulty, Samson Alemu Argaw, Shira Doron, Maya L. Nadimpalli

## Abstract

Low socioeconomic status (SES) is thought to exacerbate risks for bacterial infections, but global evidence for this relationship has not been synthesized. We systematically reviewed the literature for studies describing participants’ SES and their risk of colonization or community-acquired infection with priority bacterial pathogens. Fifty studies from 14 countries reported outcomes by participants’ education, healthcare access, income, residential crowding, SES deprivation score, urbanicity, or sanitation access. Low educational attainment, lower than average income levels, lack of healthcare access, residential crowding, and high deprivation were generally associated with higher risks of colonization or infection. There is limited research on these outcomes in low- and middle-income countries (LMICs) and conflicting findings regarding the effects of urbanicity. Only a fraction of studies investigating pathogen colonization and infection reported data stratified by participants’ SES. Future studies should report stratified data to improve understanding of the complex interplay between SES and health, especially in LMICs.

**Putting research into context:** With community-acquired antimicrobial resistance (AMR) on the rise, it is important to understand the factors that exacerbate colonization and infection with priority pathogens that are increasingly antimicrobial-resistant, particularly in the context of the social determinants of health. Previous studies have found that poverty exacerbates the risk of colonization/infection with community-acquired antimicrobial-resistant pathogens; however, other indicators of socioeconomic status (SES) including educational attainment or access to healthcare require further investigation. A comprehensive search of the scientific literature was conducted in MEDLINE (Ovid), MEDLINE Epub Ahead of Print, In-Process, In-Data-Review & Other Non-Indexed Citations, and Daily (Ovid), Global Health (Ovid), Embase (Elsevier), Cochrane Database of Systematic Reviews (Wiley), Cochrane Central Register of Controlled Trials (Wiley), and Web of Science Core Collection from inception through January 2022. All searches were based on an initial MEDLINE search developed and utilizing MeSH terminology and related keywords for the following concepts: Community-Acquired Infections, Outpatients, Ambulatory Care, Socioeconomic Factors, Health Status Disparities, Healthcare Disparities, Continental Population Groups, Ethnic Groups, Gram-Negative Bacteria, and individual ESKAPE pathogens.

**ADDED VALUE OF THIS STUDY:** This scoping review found sufficient evidence to support future systematic reviews and meta-analyses evaluating the relationship between SES and risks for colonization or infection with community-acquired bacterial pathogens that are increasingly antimicrobial-resistant. We identified 50 published papers from 14 countries reporting outcomes by participants’ education, healthcare access, income, residential crowding, SES deprivation score, urbanicity, or sanitation access. Low educational attainment, lower than average income levels, lack of healthcare access, residential crowding, and high deprivation were generally associated with higher risks of colonization and infection.

**IMPLICATIONS OF ALL THE AVAILABLE EVIDENCE:** This review identified several gaps in the current literature describing relationships between SES and risks for colonization/infection with community-acquired bacterial pathogens. First, we identified few studies from LMICs, despite LMICs having the highest burden of AMR. Only a fraction of published studies reported data stratified by SES, as SES is more often controlled for rather than analyzed as an exposure of interest in bacterial colonization and infection studies. Of the studies that did report results stratified by SES, few examined collinearity between reported SES characteristics, making it challenging to assess the most important exposures driving or mediating observed associations. Future studies should report data stratified by SES characteristics or SES deprivation scores to allow for a better understanding of the complex interplay between SES and health, especially in LMICs.

## Introduction

Antimicrobial resistance (AMR) poses a significant and urgent threat to global public health, as emphasized by the World Health Organization (1). Nearly 5 million deaths were associated with bacterial AMR in 2019 worldwide, primarily caused by the pathogens *Escherichia coli*, *Staphylococcus aureus*, *Klebsiella pneumoniae*, *Streptococcus pneumoniae*, *Acinetobacter baumannii*, and *Pseudomonas aeruginosa* (2).

Antibiotic use selects for AMR; thus, AMR is exacerbated when antibiotics are misused or overused due to lack of access to clean water, sanitation, and hygiene (WASH) for humans and animals; poor infection control in healthcare facilities and farms; poor access to quality, affordable medicines, vaccines, and diagnostics; lack of awareness and knowledge about AMR; and insufficient regulations for antibiotic purchasing (3). Conceivably, the conditions affecting where people are born, live, learn, work, play, worship, and age (4) may exacerbate the risk of acquiring AMR bacterial pathogens.

Previous studies have found that poverty exacerbates the risk of colonization/infection with community-acquired antimicrobial-resistant bacteria (8). However, other indicators of socioeconomic status (SES) related to the social determinants of health (SDOH), such as educational attainment or access to healthcare, require further investigation. This underscores the critical need for a comprehensive review of the available evidence to understand socioeconomic disparities in the risk of acquiring AMR, both in the U.S. and globally.

The overarching goal of this scoping review is to compile evidence investigating the association between SES and differential colonization/infection with AMR-bacterial pathogens. We broadly defined SES and included relevant literature from any country, provided it included data regarding colonization or community-acquired infection with one or more bacterial species commonly associated with AMR, namely *Enterococcus faecium*, *Staphylococcus aureus*, *Klebsiella pneumoniae*, *Acinetobacter baumannii*, *Pseudomonas aeruginosa*, *Enterobacter* species, and *Escherichia coli*.

## Methods

### Search strategy and selection criteria

The search strategy for this scoping review was constructed to support the current study as well as a scoping review of the evidence for racial & ethnic disparities in community-acquired colonization/infection with the 7 bacterial species of interest (5). A comprehensive search of the scientific literature was conducted in MEDLINE (Ovid), MEDLINE Epub Ahead of Print, In-Process, In-Data-Review & Other Non-Indexed Citations, and Daily (Ovid), Global Health (Ovid), Embase (Elsevier), Cochrane Database of Systematic Reviews (Wiley), Cochrane Central Register of Controlled Trials (Wiley), and Web of Science Core Collection for eligible studies that reported race, ethnicity, or SES for populations with a pathogen of interest. Search strategies were designed using a combination of controlled vocabulary and free-text keywords. All searches were based on an initial MEDLINE search developed in collaboration among the authors and utilizing MeSH terminology and related keywords for the following concepts: Community-Acquired Infections, Outpatients, Ambulatory Care, Socioeconomic Factors, Health Status Disparities, Healthcare Disparities, Continental Population Groups, Ethnic Groups, Gram-Negative Bacteria, and individual ESKAPE pathogens. The MEDLINE strategy (**Supplementary Table 1**) was translated to each of the listed databases by RM, and all databases were searched from inception through January 2022, except for MEDLINE Epub Ahead of Print, In-Process, In-Data-Review & Other Non-Indexed Citations and Daily for which the search covered 2017 through 10 January 2021. References were collected and deduplicated using Endnote X9 before export to Covidence (6) for screening and management.

### Eligibility Criteria

We included studies that reported data on SES for at least one of the pathogens of interest irrespective of study designs (except case-control or case series), age group, or country. We included studies reporting both infection and colonization, as long as data were reported separately. We excluded studies reporting mixed pathogens if more than 50% of the pathogens were not of interest unless they reported subgroup data for at least one of our pathogens of interest. Studies had to specify that the pathogen of interest was community-acquired or report only outpatient or community-based data. Studies that did not report outpatient or community data and which defined community acquisition based only on phenotype (e.g., susceptibility to gentamicin for *Staphylococcus aureu*s) or sequence type (e.g., USA300 for *Staphylococcus aureu*s), were excluded. In studies that included both hospital-acquired and community-acquired data, we excluded studies that did not report subgroup data for the community-acquired pathogen. In studies reporting colonization, we excluded those that compared persistent colonization to cleared colonization. We also excluded studies that reported results of comparisons between countries, regions, or hospitals, rather than persons.

### Screening, Data Extraction, and Synthesis

Abstracts identified from the literature searches and full-text articles of eligible citations were accessed using the predefined inclusion and exclusion criteria (**Table 1**). Full-text articles were exported if their abstract suggested they met inclusion criteria and then screened further.

**Table 1.** Exclusion and Inclusion Criteria.

|  | Inclusion Criteria | Exclusion Criteria |
| --- | --- | --- |
| Study design | Observational studies | Case reports/study<br>Case series<br>Narrative Review |
| Study population | Any age group, gender, country, and health status | Studies in which data for race or ethnicity or socioeconomic factors were not reported |
| Exposure | Any socioeconomic factor reported | Data stratified by country, region, or hospital rather than a racial/ethnic group or socioeconomic status |
| Comparator | Distinct socioeconomic status group from the same country | No distinct socioeconomic metrics reported |
| Outcomes | Community-acquired (e.g. outpatient, ERs, health clinics, ambulatory care, population-level surveillance) colonization or infection with:<br>▪ *Enterococcus faecium*<br>▪ *Staphylococcus aureus*<br>▪ *Klebsiella pneumoniae*<br>▪ *Acinetobacter baumannii*<br>▪ *Pseudomonas aeruginosa*,<br>▪ *Enterobacter* species<br>▪ *Escherichia coli*<br>▪ Enterobacteriaceae or Enterobacterales | Hospital-acquired colonization or infection<br>Device-associated (e.g. catheter) infection<br>Non-bacterial infection or cause of infection unclear<br>Mixed pathogen infections if less than 50% of the pathogens were of interest<br>Community-acquisition or community-association based only on phenotype or genotype |

Citations and full-text articles were screened independently by two reviewers and any conflicts were resolved in consensus during weekly team meetings. Covidence was used for abstract and full-text screening, data extraction, and data management. A customized extraction form was created in Covidence (6) to capture relevant study data from eligible studies including study design, study definition of community-acquired, study population characteristics, exposure and comparator, outcomes of interest, and directionality of results. For each paper, we extracted all possible comparisons between an SES exposure (e.g. healthcare access) and an outcome (e.g. methicillin-resistant *Staphylococcus aureus* (MRSA) infection) regardless of whether the original authors conducted a statistical test to assess the significance of that association. We refer to each of these extracted exposure-outcome pairs as “comparisons” in this manuscript. We note that in situations where a study reported data stratified by more than one SES exposure and/or more than one outcome of interest, we extracted multiple comparisons from that study. The extraction form was piloted by the author team for a subset of studies and then revised to ensure that all relevant data were captured. Each study was extracted independently by two team members into the standard extraction form. Another team member compared the data entries of the two extractors and resolved any discrepancies. Extracted data from all included studies are summarized in tables and figures using R version 4.3.1 (7) and ggplot2 package (8).

## Results

Our literature search retrieved 1039 unique citations, 388 of which met preliminary inclusion criteria. After full-text screening of these 388 articles, 85 continued to meet the inclusion criteria (**Figure 1**). Two additional articles were identified by searching the paper’s references, contributing to a total of 87 papers included. Fifty of these full-text articles reported colonization/infection with one or more of the seven bacterial species of interest by participants’ SES. **Supplementary Table 2** describes the 50 studies included in this scoping review, including country and date of publication, recruitment method, bacterial species, study population demographics, and clinical diagnosis (e.g. SSTI, UTI), if reported. Studies from 14 countries were included (**Figure 2**). Thirty-seven papers reported data from high-income countries, two from upper-middle-income countries, eight from lower-middle-income countries (LMICs), and three from low-income countries. Studies were published from 1992 and onwards (**Figure 3**). Each study reported data stratified by at least one of seven indicators of SES: education (n=13 studies), healthcare access (n=14), income (n=18), residential crowding (n=16), SES deprivation score (n=14), urbanicity (n=12 papers), and water, sanitation, and hygiene (WASH) access (n=4). Most reports were among participants experiencing colonization/infection with *S. aureus* (**Figure 4**).

**Figure 1.**
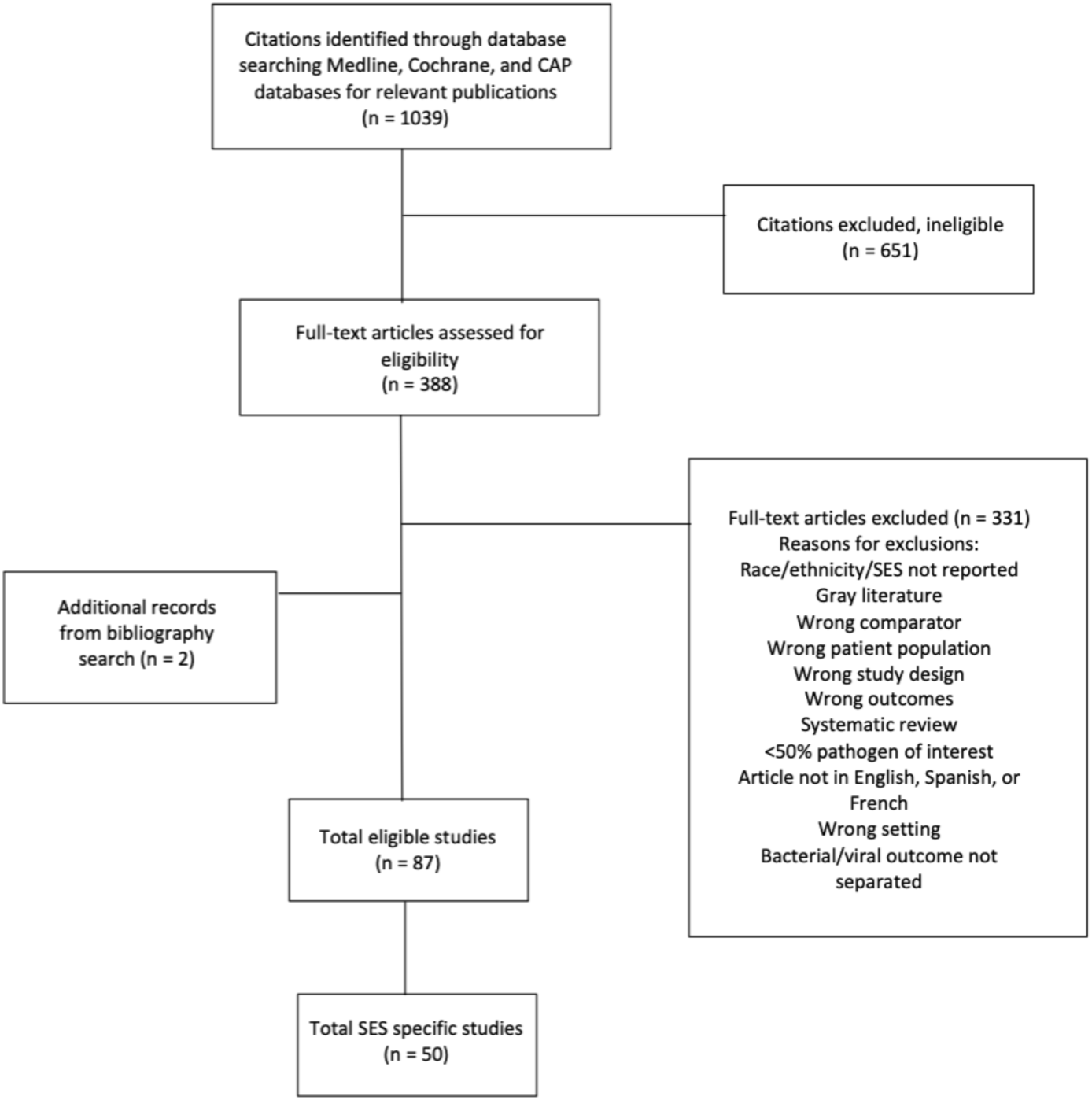
Study inclusion schema.

**Figure 2.**
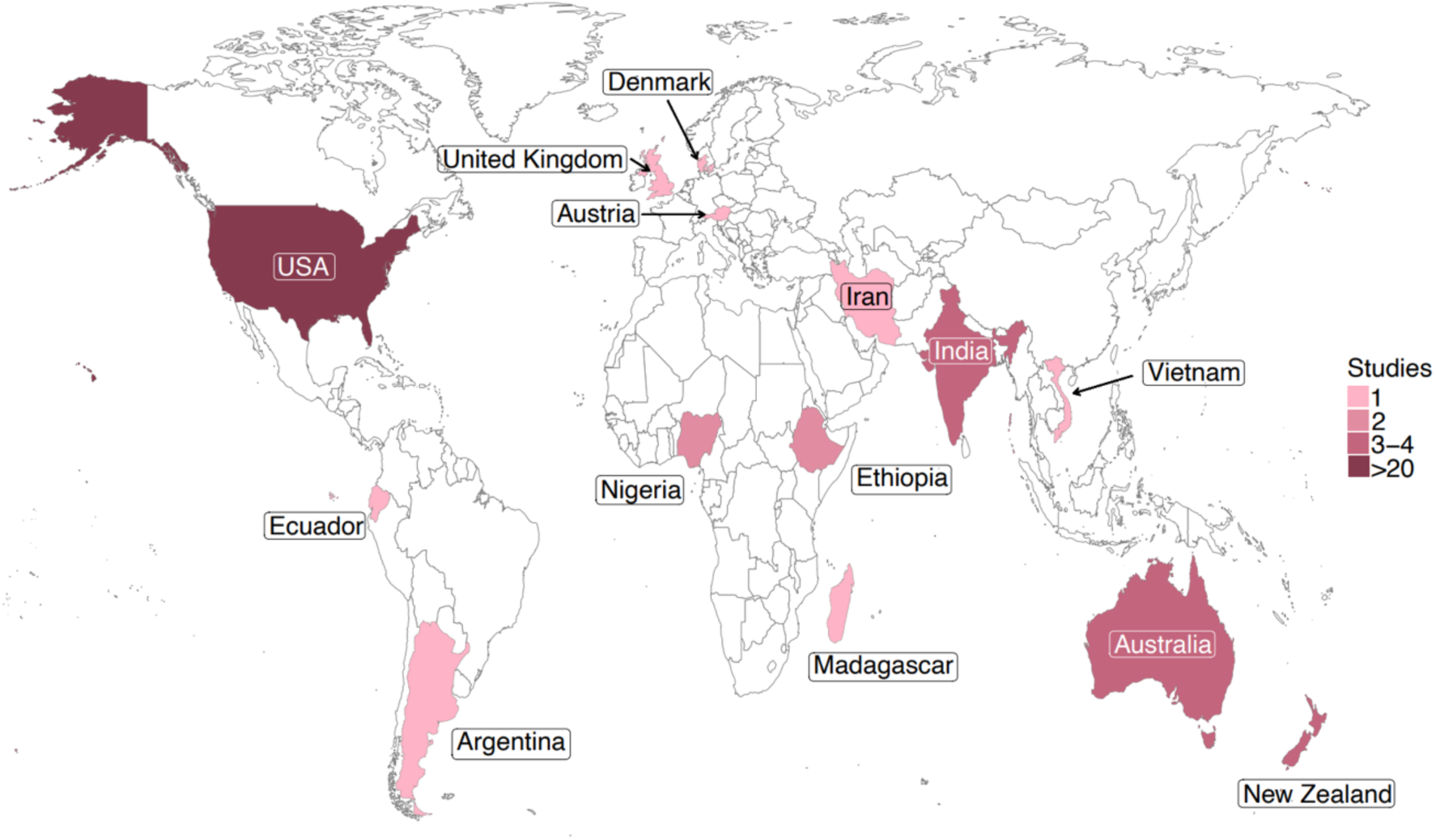
Study location for 50 studies describing individuals’ risk of colonization or infection with priority pathogens by their socioeconomic status.

**Figure 3:**
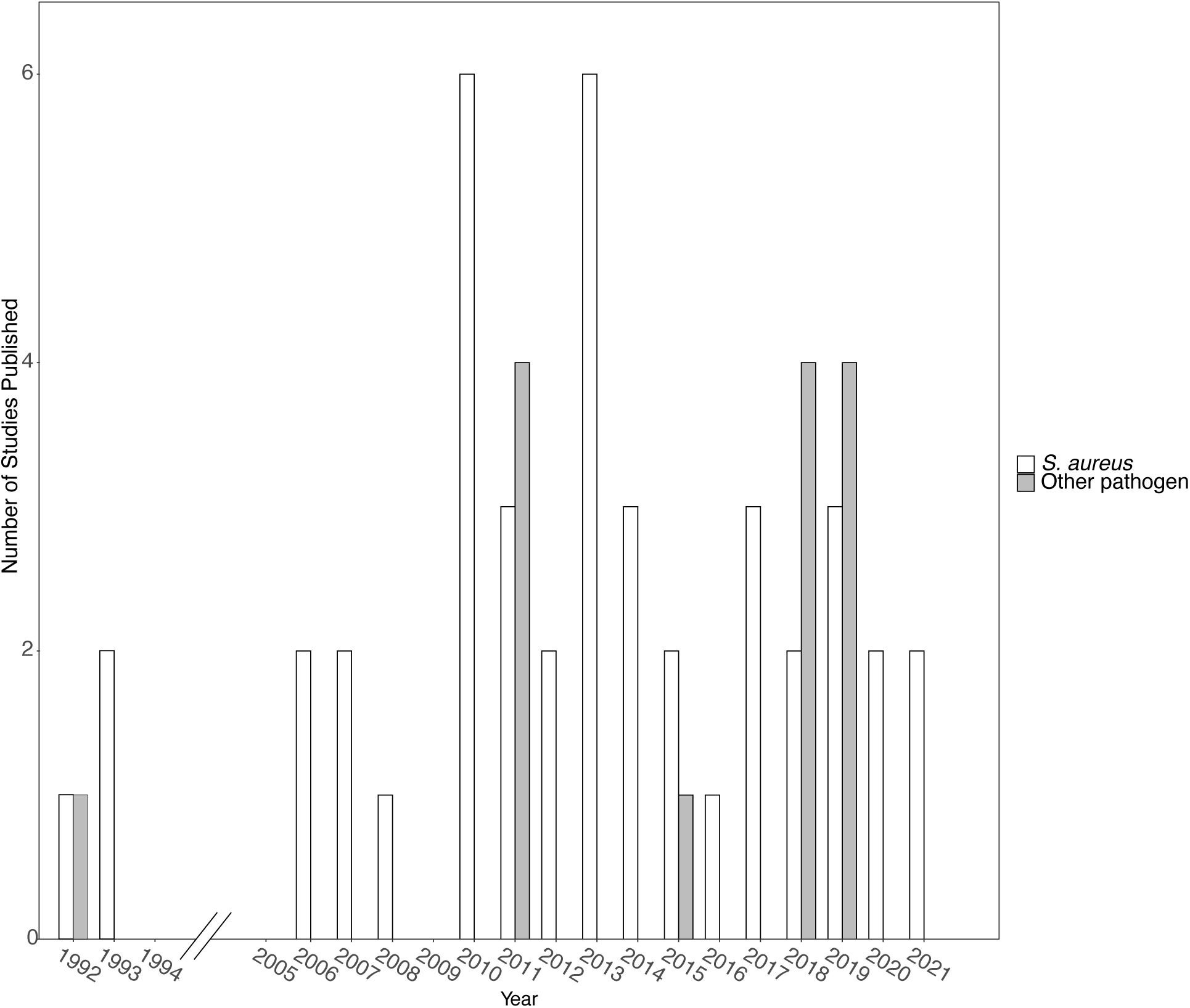
Year of publication for 50 included studies.

**Figure 4.**
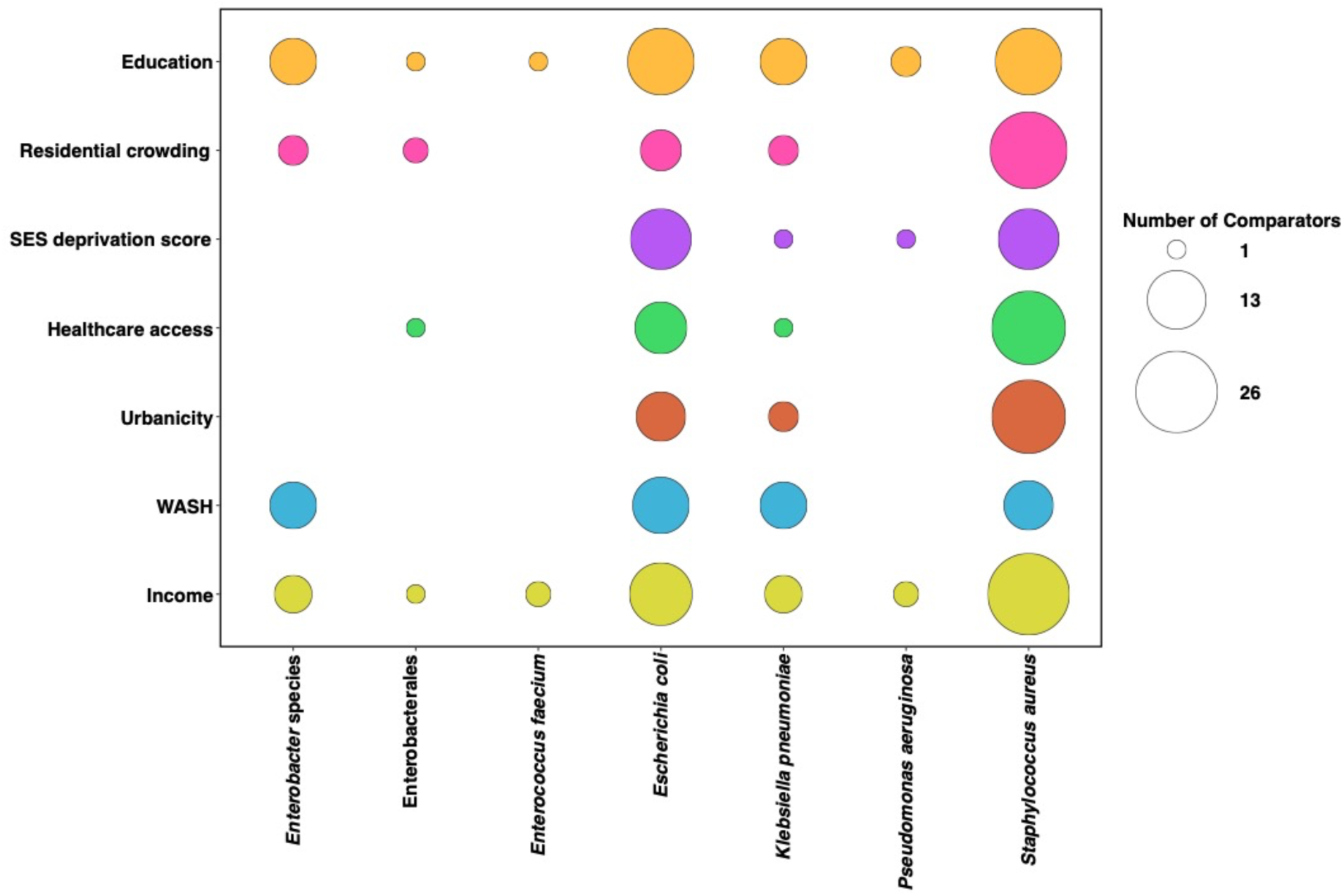
Distribution of indicators of socioeconomic status by 7 priority pathogens among 50 publications included in this scoping review. *<u>Note</u>*: Comparison was counted more than once for visualization purposes, if the outcome reported (i.e., SSTI, UTI, bacteremia, bacteriuria) was caused by more than one organism of interest.

### Education

Thirteen studies reported participants’ education status (9–21) (**Table 2**). We extracted one to five comparisons from each (n=31 total). Among the 25 comparisons for which authors evaluated the association between participants’ educational attainment and an outcome of interest, 13 were associated with lower educational attainment. Shorter duration of education was significantly associated with a greater risk of *K. pneumoniae* oropharyngeal tract colonization among community-dwelling adults and children in Vietnam (11) and a higher incidence of *S. aureus* bacteremia, *E. coli* bacteremia, and community-acquired bacteremia (CAB) among Danish patients (15). One paper reported that compared to participants who had a college-level education or higher, having no formal education was associated with a greater risk of bacteriuria among adults visiting an outpatient department in Ethiopia (17). Seven comparisons from five studies found that relative to individuals who had completed more than high school, lower educational attainment was associated with a significantly higher risk of gut colonization with multidrug-resistant (MDR) and extensively drug-resistant *E. coli* among children in Ecuador (16), MRSA infections among U.S. patients (10,20,21), and nitrofurantoin-resistant *E. coli* infections among UK patients (18). One paper reported that, compared to the head of a family being a manager, pregnant women whose head of a family was unemployed, unskilled, semi-skilled, or a non-manual employee had a higher risk of intestinal colonization with extended-spectrum beta-lactamase (ESBL)-producing Enterobacterales (14). Twelve comparisons from eight studies did not find a statistically significant association between participants’ education status and their risk of colonization/infection with an outcome of interest (9,10,12,14–16,18,19).

**Table 2.** Characteristics of publications reporting colonization or infection with a pathogen of interest by subject’s level of educational attainment.

|  | Total (%) |
| --- | --- |
| **N studies** | 13 |
| **Age Group** |  |
| Children Only | 3 (23%) |
| Adults Only | 5 (38%) |
| Multiple age groups<sup>a</sup> | 4 (31%) |
| Not Reported | 1 (8%) |
| **Study length** | 5 months – 8 years |
| **Country** |  |
| Denmark | 1 (8%) |
| Ecuador | 1 (8%) |
| Ethiopia | 2 (15%) |
| India | 3 (23%) |
| Madagascar | 1 (8%) |
| United Kingdom | 1 (8%) |
| United States | 3 (23%) |
| Vietnam | 1 (8%) |
| **Outcome type** |  |
| Colonization | 5 (38%) |
| Infection | 8 (62%) |
| **Data source** |  |
| Inpatient recruitment | - |
| Outpatient recruitment | 6 (46%) |
| Patient medical records | 4 (31%) |
| Community-based recruitment | 1 (8%) |
| Household-based recruitment | 2 (15%) |
| Multiple | - |
| **Pathogen<sup>b</sup>** |  |
| *Staphylococcus aureus* | 9 (30%) |
| *Enterococcus faecium* | 1 (3%) |
| *Escherichia coli* | 8 (27%) |
| *Pseudomonas aeruginosa* | 3 (10%) |
| *Klebsiella pneumoniae* | 3 (10%) |
| *Enterobacter* species | 5 (17%) |
| *Acinetobacter baumannii* | 1 (3%) |

<sup>a</sup>i.e., adults, seniors, and/or children
<sup>b</sup>Denominators for these percentages is 30 not 13 because publications looked at more than one pathogen species

### Healthcare Access

Fourteen U.S. studies reported outcomes of interest by participants’ healthcare access (13,21–33) (**Table 3**). We extracted up to eight comparisons from each (n=32 total). Among the 21 comparisons for which authors evaluated the association between healthcare access and an outcome of interest, more than 75% of comparisons were found to have a statistically significant association. One study found that compared to using private or military insurance, having Medicaid, Medicare, or no health insurance was associated with a significantly elevated risk of *S. aureus* colonization (33). A U.S. study surveyed medical records from two healthcare facilities on the West Coast and found that Medicaid use was associated with a 0.6% and 0.9% reduction in the risk of being diagnosed with a urinary tract infection (UTI). They also found that Medicaid use was associated with an 8-9% increase in the risk of UTI caused by MDR *E. coli* (28). Two studies found that children with Medicaid or no insurance were more likely to be colonized (24) or infected with MRSA (26) than children with private insurance or commercial plans. Similar trends have been reported in adults; five papers found adults relying on public insurance (22,25,31), self-pay (30,31), or other insurance (30) had a significantly higher risk of MRSA infection than those using private insurance. A national study found that the risk of MRSA infection among those living in a medically underserved area was 2.4 times the risk among those not living in a medically underserved area (21). Additionally, the researchers found that the risk of MRSA infection was 92% lower for those with healthcare coverage compared to those with no healthcare coverage (21). One study found that requiring an interpreter was associated with a 28 to 36% increase in the risk of UTI caused by MDR *E. coli* (28). A northwest U.S. study found that compared to those with insurance, uninsured patients of a county Hospital system were at a greater risk for infections with ceftriaxone-resistant Enterobacteriaceae and ceftriaxone-resistant *E. coli* (23). Three comparisons from three studies did not find a statistically significant association between participants’ healthcare access and their risk of colonization/infection with an outcome of interest (27,29,32).

**Table 3.** Characteristics of publications reporting colonization or infection with a pathogen of interest by subject’s access to healthcare.

|  | Total (%) |
| --- | --- |
| **N studies** | 14 |
| **Age Group** |  |
| Pediatric Only | 7 (50%) |
| Adults Only | 4 (29%) |
| Mixed age groups<sup>a</sup> | 3 (21%) |
| Not Reported | - |
| **Study duration range** | 8 months – 12 years |
| **Study Country** |  |
| United States | 14 (100%) |
| **Carriage type** |  |
| Colonization | 4 (29%) |
| Infection | 10 (71%) |
| **Recruitment setting** |  |
| Inpatients | 3 (21%) |
| Outpatients | 2 (14%) |
| Medical records | 7 (50%) |
| Community | 2 (14%) |
| Household | - |
| Mixed | - |
| **Pathogen<sup>b</sup>** |  |
| *Staphylococcus aureus* | 11 (61%) |
| *Enterococcus fecium* | - |
| *Escherichia coli* | 3 (17%) |
| *Pseudomonas aeruginosa* | - |
| *Klebsiella pneumoniae* | 2 (11%) |
| Enterobacterales | 1 (6%) |
| *Enterobacter* species | 1 (6%) |
| *Acinetobacter baumannii* | - |

<sup>a</sup>*i.e.*, adults, seniors, and/or children
<sup>b</sup>Denominators for these percentages is 18 not 14 because publications looked at more than one pathogen species

### Income

Eighteen studies reported outcomes of interest by participants’ income status (9–12,15,16,18,20–23,25,31,34–38) (**Table 4**). Among the 36 comparisons for which authors statistically evaluated the association between participants’ income and an outcome of interest, approximately nine comparisons demonstrated an association between low-income levels and elevated risk of colonization/infection with priority pathogens, regardless of their susceptibility to antibiotics. Four comparisons from a Danish study found having a low personal annual income was associated with a significantly elevated risk for CAB and bacteremia caused by *S. aureus*, *Enterococci*, or *E. coli* (15). An Ethiopian study found that children with a caretaker who had a low monthly income (less than 1000 Ethiopian Birr) were more likely to be gut-colonized with diarrheagenic *E. coli* (12). A study conducted on the West Coast of the U.S. found that patients living in neighborhoods with the lowest quintile of income had 17% higher odds of *S. aureus* SSTI (versus other causes of SSTI) than patients living in neighborhoods with the highest quintile of income (38). An Indian study found that pregnant women in the lower half of the household income distribution were at greater risk for bacteriuria (9). They also found that women in the bottom half of the household income distribution had a greater risk of having bacteriuria caused by ESBL-producing organisms and being colonized with ESBL bacteria (9).

**Table 4.** Characteristics of publications reporting colonization or infection with a pathogen of interest by subject’s level of income.

|  | Total (%) |
| --- | --- |
| **N studies** | 18 |
| **Age Group** |  |
| Pediatric Only | 5 (28%) |
| Adults Only | 3 (17%) |
| Mixed age groups<sup>a</sup> | 9 (50%) |
| Not Reported | 1 (6%) |
| **Study duration range** | 5 months – 9 years |
| **Study Country** |  |
| Denmark | 1 (6%) |
| Ecuador | 1 (6%) |
| Ethiopia | 1 (6%) |
| India | 3 (17%) |
| United Kingdom | 1 (6%) |
| United States | 10 (56%) |
| Vietnam | 1 (6%) |
| **Carriage type** |  |
| Colonization | 6 (33%) |
| Infection | 12 (67%) |
| **Recruitment setting** |  |
| Inpatients | - |
| Outpatients | 3 (17%) |
| Medical records | 11 (61%) |
| Community | 1 (6%) |
| Household | 2 (11%) |
| Mixed | 1 (6%) |
| **Pathogen<sup>b</sup>** |  |
| *Staphylococcus aureus* | 14 (44%) |
| *Enterococcus fecium* | 1 (3%) |
| *Escherichia coli* | 7 (22%) |
| *Pseudomonas aeruginosa* | 2 (6%) |
| *Klebsiella pneumoniae* | 2 (6%) |
| Enterobacterales | 1 (3%) |
| *Enterobacter* species | 4 (16%) |
| *Acinetobacter baumannii* | 1 (3%) |

<sup>a</sup>i.e., adults, seniors, and/or children
<sup>b</sup>Denominators for these percentages is 32 not 18 because publications looked at more than one pathogen species

Eleven additional comparisons identified an association between low income and elevated risk of colonization/infection with antibiotic-resistant pathogens. Three U.S. studies found that low-income households had an overall increased risk of MRSA infection (10,21,22), as well as an increased risk of having MRSA over methicillin-sensitive *S. aureus* (MSSA) (10). Researchers concluded that persons residing in low-income neighborhoods on the West Coast of the U.S. were 1.43 times more likely to have MRSA SSTI compared to MSSA SSTI (38). A national study found that patients living in poverty in the past 12 months had 16.78 times the risk of invasive MRSA infection versus no MRSA compared to patients who had not reported living in poverty (21). They also found that patients living in neighborhoods with a higher percentage of homes valued at ≥400% the median home value (i.e., $750,000) had half the risk of MRSA infection than their counterparts (21). Further, patients living in neighborhoods with extreme income inequality had nearly 13 times greater risk of MRSA infection than patients living in neighborhoods with less income inequality (21). Two studies from the southeast U.S. reported that children living in a block group where the majority of households lived below the poverty level had higher odds of community-onset MRSA SSTI (25) and were at greater risk of MRSA SSTI relative to MSSA SSTI (31). One Midwest U.S. study found similar results that low-income individuals were at greater risk for MSSA infections relative to non-MSSA infections (10). Overall, there were a total of 16 comparisons where the authors did not find a statistically significant association between participants’ income status and their risk of colonization/infection with an outcome of interest (11,15,16,18,20,23,34,36,37).

### Residential Crowding

Sixteen studies reported outcomes of interest by participants’ residential crowding conditions (10,14,16,21,23–25,31,39–46) (**Table 5**). We extracted between one to three comparisons from each study (n=29 total). Among the 27 comparisons for which authors evaluated the association between participants’ residential crowding status and an outcome of interest, two comparisons found that having more than two household occupants in a bedroom was associated with an increased risk of *S. aureus* colonization for children living in New Zealand (42). Another study in Argentina found that living in homes with greater than 3 persons per room was associated with an increased risk for *S. aureus* SSTI (39). Four comparisons found that having more than 1 person per room was associated with an increased risk of MRSA infection (31), SSTI (including MRSA, MSSA, and other pathogens) (40), or ceftriaxone-resistant *E. coli* (23). Two more comparisons found that having more than 2 persons per bedroom (24) and an increasing ratio of inhabitants per room (14) was associated with an increased risk of MRSA colonization in children from Madagascar and ESBL carriage in adults from Madagascar, respectively. Three comparisons from the U.S. found that repeat (46), recent (43), or history of incarceration (44) were associated with an increased risk of MRSA colonization and MRSA SSTI in adults. One paper found that compared to those living in stable housing, those in temporary housing were at an increased risk of MRSA colonization (43). Two comparisons found that the odds of MRSA infection for current residence in public housing (43) or having lived in a group setting (10) was 2.5 to 3.9 times the odds of MRSA infection among those not residing in public housing or ever lived in a group setting, respectively. There were a total of 11 comparisons where the authors did not find a statistically significant association between participants’ residential crowding condition and their risk of colonization/infection with an outcome of interest (10,16,23,41–45).

**Table 5.** Characteristics of publications reporting colonization or infection with a pathogen of interest by subject’s level of residential crowding.

|  | Total (%) |
| --- | --- |
| **N studies** | 16 |
| **Age Group** |  |
| Pediatric Only | 5 (31%) |
| Adults Only | 7 (44%) |
| Mixed age groups<sup>a</sup> | 4 (25%) |
| Not Reported | - |
| **Study duration range** | 37 days – 9 years |
| **Study Country** |  |
| Argentina | 1 (6%) |
| Ecuador | 1 (6%) |
| Madagascar | 1 (6%) |
| New Zealand | 1 (6%) |
| United States | 12 (75%) |
| **Carriage type** |  |
| Colonization | 6 (38%) |
| Infection | 10 (63%) |
| **Recruitment setting** |  |
| Inpatients | - |
| Outpatients | 4 (25%) |
| Medical records | 8 (50%) |
| Community | 1 (6%) |
| Household | 2 (13%) |
| Mixed | 1 (6%) |
| **Pathogen<sup>b</sup>** |  |
| *Staphylococcus aureus* | 13 (62%) |
| *Enterococcus fecium* | - |
| *Escherichia coli* | 3 (14%) |
| *Pseudomonas aeruginosa* | - |
| *Klebsiella pneumoniae* | 2 (10%) |
| Enterobacterales | 1 (5%) |
| *Enterobacter* species | 2 (10%) |
| *Acinetobacter baumannii* | - |

<sup>a</sup>i.e., adults, seniors, and/or children
<sup>b</sup>Denominators for these percentages is 21 not 16 because publications looked at more than one pathogen species

### SES Deprivation Score

Thirteen relevant studies were identified (18,20,28,39,42,44,47–53) (**Table 6**). We extracted between one to eight comparisons from each (n=27 total). Among the 25 comparisons for which authors evaluated the association between SES deprivation score and an outcome of interest, four comparisons from New Zealand found that medium to high deprivation deciles were associated with an increased risk of pediatric *S. aureus* or MRSA infection (42,51,53). One comparison found that participants living in U.S. communities above the median community SES deprivation score (based on six indicators derived from U.S. Census 2000 data) (47) was associated with a risk of *S. aureus* SSTI. Another comparison found an increased risk of *S. aureus* SSTI was associated with those living in neighborhoods in Argentina with more deprivation (unsatisfied basic needs type 3) (39). One study utilized the 2016 Index of Relative Social Economic Disadvantage [IRSD] to assess SES deprivation and ceftriaxone-resistant *E. coli* uropathogens in Victoria, Australia, and found that living in a community ranked as 1st decile (most deprived) was associated with an increased risk for this outcome (48). Eight comparisons found that compared to those living in low-deprivation neighborhoods, those living in neighborhoods with high SES deprivation scores had an increased risk of infection with MRSA (22,47), MDR *E. coli* (28), trimethoprim-resistant *E. coli* (18), ampicillin-resistant *E. coli* (18), ciprofloxacin-resistant *E. coli* (18), and nitrofurantoin-resistant *E. coli* (18). There were a total of 10 comparisons where the authors did not find a statistically significant association between participants’ SES deprivation score and their risk of colonization/infection with an outcome of interest (18,20,42,44,49,50,52).

**Table 6.** Characteristics of publications reporting colonization or infection with a pathogen of interest by subject’s SES Deprivation Score.

|  | Total (%) |
| --- | --- |
| **N studies** | 14 |
| **Age Group** |  |
| Pediatric Only | 4 (29%) |
| Adults Only | 4 (29%) |
| Mixed age groups<sup>a</sup> | 5 (36%) |
| Not Reported | 1 (7%) |
| **Study duration range** | 1 month – 9 years |
| **Study Country** |  |
| Argentina | 1 (7%) |
| Australia | 3 (21%) |
| India | 1 (7%) |
| New Zealand | 3 (21%) |
| Nigeria | 1 (7%) |
| United Kingdom | 1 (7%) |
| United States | 4 (29%) |
| **Carriage type** |  |
| Colonization | 2 (14%) |
| Infection | 12 (86%) |
| **Recruitment setting** |  |
| Inpatients | - |
| Outpatients | 3 (21%) |
| Medical records | 8 (57%) |
| Community | 1 (7%) |
| Household | - |
| Mixed | 2 (14%) |
| **Pathogen<sup>b</sup>** |  |
| *Staphylococcus aureus* | 11 (52%) |
| *Enterococcus fecium* | - |
| *Escherichia coli* | 5 (24%) |
| *Pseudomonas aeruginosa* | 2 (10%) |
| *Klebsiella pneumoniae* | 1 (5%) |
| Enterobacterales | - |
| *Enterobacter* species | 1 (5%) |
| *Acinetobacter baumannii* | 1 (5%) |

<sup>a</sup>i.e., adults, seniors, and/or children
<sup>b</sup>Denominators for these percentages is 21 not 14 because publications looked at more than one pathogen species

### Urbanicity

Twelve studies from eight countries reported outcomes of interest by participants’ urbanicity status (11,18,21,22,28,42,47,48,54–57) (**Table 7**). We extracted between one to five comparisons from each study (n=33 total). Among the 21 comparisons for which authors evaluated the association between participants’ urbanicity status and an outcome of interest, most found that living in a non-urban setting was associated with an increased risk for colonization/infection with an outcome of interest (11,22,56). However, the opposite relationship was also reported (21,47). Two comparisons found that compared to living in an urban location, those living in a rural location had an increased risk of oropharyngeal *K. pneumoniae* carriage (11) and *S. aureus* colonization (56). One paper found that Vietnamese community members employed as farmers or hired laborers had a greater risk of oropharyngeal *K. pneumoniae* carriage (11). One comparison found that patients from central and northeastern Pennsylvania living in a city, rather than a township, had increased odds of *S.aureus* SSTI (47). In the northwest US, residents with greater proximity to swine farms were at greater risk of MRSA SSTI versus No MRSA SSTI (22). An Austrian study found that living in a rural setting was associated with a greater risk of being colonized with resistant *S. aureus* (56). On the other hand, two comparisons found that living in a U.S. city (47) or a U.S. community with high urbanicity (21) was associated with an increased risk of MRSA infection. There were a total of 11 comparisons where the authors did not find a statistically significant association between participants’ urbanicity status and their risk of colonization/infection with an outcome of interest (18,42,48,56).

**Table 7.**
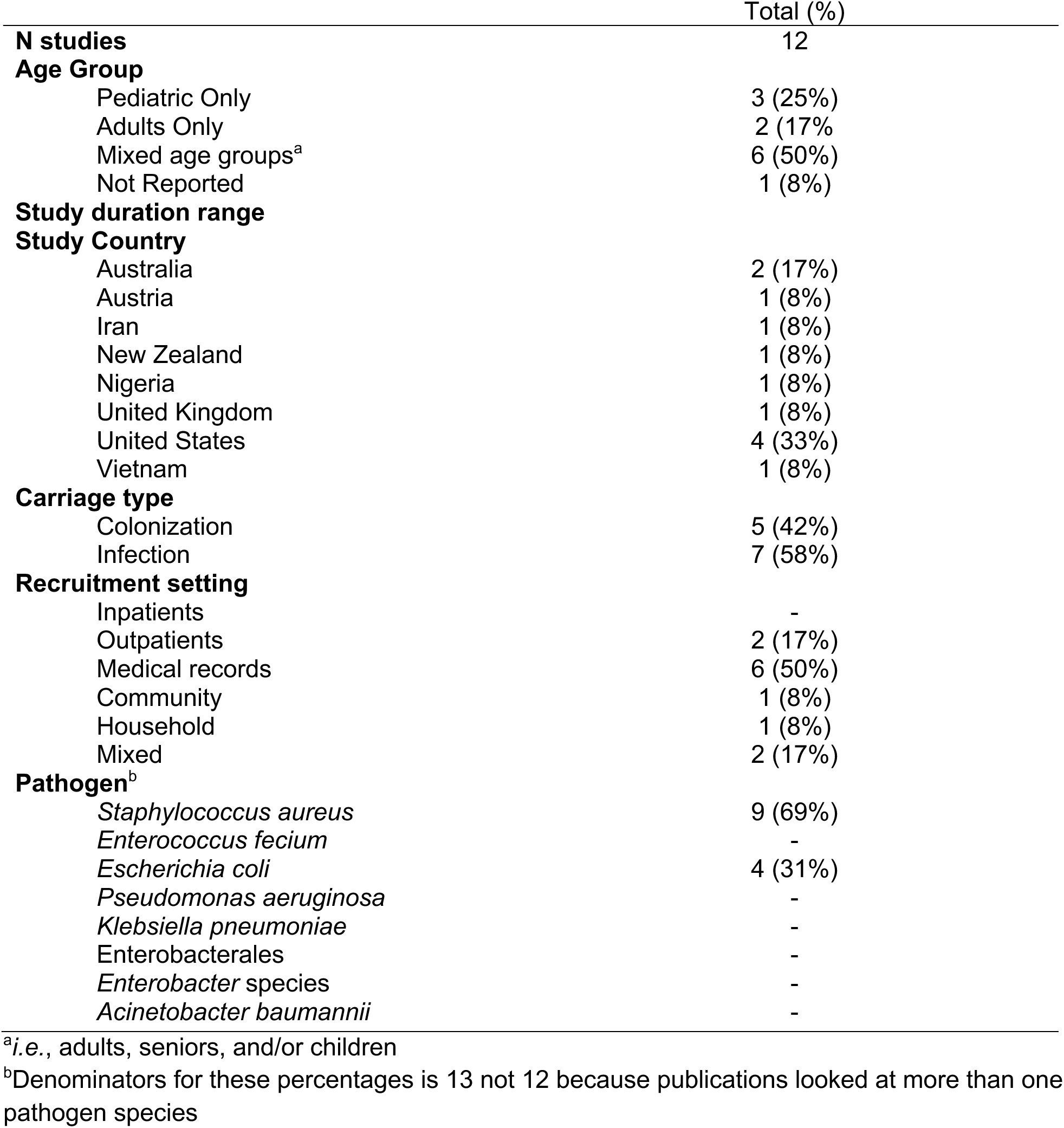
Characteristics of publications reporting colonization or infection with a pathogen of interest by subject’s level of urbanicity.

### WASH

Four studies reported outcomes of interest by participants’ WASH access (9,12,16,58) (**Table 8**). We extracted up to 8 comparisons from each (n=13 total). For six comparisons, the authors did not investigate the association between WASH and an outcome of interest (9). Crum-Cianflone 2011 found that using a public bath was associated with 6.91 times the odds of MRSA colonization among HIV-infected U.S. adults relative to no public bath use (58). No other comparisons were statistically significant (9,12,16).

**Table 8.** Characteristics of publications reporting colonization or infection with a pathogen of interest by subject’s access to water, sanitation, and hygiene (WASH).

|  | Total (%) |
| --- | --- |
| **N studies** | 4 |
| **Age Group** |  |
| Pediatric Only | 2 (50%) |
| Adults Only | 2 (50%) |
| Mixed age groups<sup>a</sup> | - |
| Not Reported | - |
| **Study duration range** | 5 months – 3 years |
| **Study Country** |  |
| Ecuador | 1 (25%) |
| Ethiopia | 1 (25%) |
| India | 1 (25%) |
| United States | 1 (25%) |
| **Carriage type** |  |
| Colonization | 3 (75%) |
| Infection | 1 (25%) |
| **Recruitment setting** |  |
| Inpatients | - |
| Outpatients | 3 (75%) |
| Medical records | - |
| Community | - |
| Household | 1 (25%) |
| Mixed | - |
| **Pathogen<sup>b</sup>** |  |
| *Staphylococcus aureus* | 1 (14%) |
| *Enterococcus fecium* | - |
| *Escherichia coli* | 3 (43%) |
| *Pseudomonas aeruginosa* | - |
| *Klebsiella pneumoniae* | 2 (29%) |
| Enterobacterales | - |
| *Enterobacter* species | 1 (14%) |
| *Acinetobacter baumannii* | - |

<sup>a</sup>*i.e.*, adults, seniors, and/or children
<sup>b</sup>Denominators for these percentages is 7 not 4 because publications looked at more than one pathogen species

## Discussion

It is well-established that the global burden of bacterial infections differs among countries. However, whether individual susceptibility might vary within countries, particularly as a function of the social determinants of health, has not been comprehensively examined. Such risk factors are important to establish given that many bacterial infections are now increasingly challenging to treat. In this scoping review, we systematically extracted evidence from published studies that an individual’s SES may be associated with their risk of colonization/infection with seven bacterial pathogens that are frequently antibiotic-resistant, including *E. faecium*, *S. aureus*, *K. pneumoniae*, *A. baumannii*, *P. aeruginosa*, *Enterobacter* spp, and *E. coli*. Among the 50 studies published that met inclusion criteria, we noted that individuals’ risk for colonization/infection was typically associated with low educational attainment, low-income levels, lack of healthcare access, residential crowding, and high neighborhood-level deprivation. Evidence for associations with urbanicity was mixed. Notably, we identified few studies from LMICs where the burden of AMR is highest. Overall, this scoping review suggests that the social determinants of health are likely important yet understudied factors shaping individuals’ risk for being colonized or infected with bacterial pathogens that are increasingly antibiotic-resistant.

Income was one of the most reported SES characteristics and was described in studies from high-income (US, New Zealand, United Kingdom), upper-middle-income (Ecuador), lower-middle-income (India, Vietnam), and low-income countries (Ethiopia). Twenty of the 36 comparisons we extracted found that low income - whether reported at the individual, household, or neighborhood level - was significantly associated with a higher risk of colonization/infection with priority bacterial pathogens, including drug-resistant *S. aureus*, *K. pneumoniae*, *E. coli*, and *Enterobacter* spp. The relationship between income and AMR is well-established at the country level (59) but in this review, we noted associations between the outcomes of interest and individual incomes among studies published in India, the United States, Ethiopia, and Denmark - countries with starkly different gross national incomes (GNI). Regardless of a country’s GNI, low-income individuals may have less access to timely medical care (30) may be more likely to use unprescribed antibiotics (*i.e.*, from friends, relatives, online, or from outside the country), (60) may live in crowded homes with poorer infrastructure (61) or lack access to hygiene, among other factors that could exacerbate their susceptibility to bacterial infection.

The only country that reported healthcare access, which we considered to be a proxy for SES, was the U.S. where insurance is not universally guaranteed and is instead typically provided through employers, resulting in profound disparities in health and healthcare access across different income groups (62). These studies defined poor healthcare access as lack of health insurance, use of public insurance, requiring a medical interpreter, or residing in a medically underserved area. Most comparisons we extracted found that indicators of poor healthcare access were associated with a higher risk of infection/colonization with a bacterial species of interest, including MRSA and drug-resistant *E. coli* causing UTIs. However, two of the 21 comparisons found the inverse relationship, where Medicaid use (i.e., public insurance for low-income persons) was associated with reduced risk of infection (28). These conflicting findings are challenging to interpret because while they could reflect true differences in susceptibility to infection, they could also reflect the fact that persons with low healthcare access may be less likely to seek treatment and be diagnosed with infections (63,64). Previous studies of pediatric patients have suggested that inadequate access to timely care for infections among uninsured patients or patients on Medicaid is a likely cause of elevated risk for resistant infections (25,29,59).

Our findings that lower education attainment was typically associated with an increased risk of colonization/infection with priority bacterial pathogens contrast that of a previous analysis of anthropological factors related to global AMR. In a country-level analysis, educational attainment may have been correlated with a population’s general access to resources (hospitals, antibiotics) (65) that could increase a population’s exposure to AMR bacteria. At an individual level, persons with less education often lack access to the information needed to make informed health decisions, leading to riskier behaviors that can increase the risk of bacterial infections (65). Our findings are more in line with general trends between educational attainment and a variety of poor health outcomes (66).

We observed conflicting trends for the association between urbanicity and an individual’s risk of colonization/infection with priority bacterial pathogens. Of the 21 comparisons we extracted from eight studies conducted in the US, Austria, New Zealand, Australia, the United Kingdom, Iran, Nigeria, and Vietnam, seven comparisons from three studies found that living in a rural setting was associated with increased risks while three comparisons from two studies reported the opposite relationship. These conflicting patterns could be explained in part by important differences in the types of exposures that may be present in some rural settings versus others. Specifically, individuals in some rural settings may be exposed to livestock or intensive animal farming which are known sources of exposure to drug-resistant *S. aureus* and *E. coli* (67) while individuals living in rural settings without frequent livestock exposures may be at lower risk of bacterial disease, compared to their urban counterparts, e.g., due to lower population density.

Further research is needed to disentangle the effects of population density, healthcare access, and exposure to livestock from residence in urban versus rural settings concerning the outcomes of interest.

More than half of the 25 comparisons we extracted from studies in seven countries found that living in a high-deprivation neighborhood was associated with an elevated risk of colonization/infection with priority bacterial pathogens, including MRSA and drug-resistant *E coli*. SES deprivation scores were first developed in the 1970s and have largely been used by high-income countries to study relationships between SES and health outcomes (68). The positive correlation between SES deprivation and risk of colonization/infection mirrors trends we observed for individual components of SES deprivation scores, like income, education, and crowding. Only one middle-income and no low-income countries stratified rates of colonization/infection with the pathogens of interest by an SES deprivation score. Deprivation scores may be more difficult to calculate in some LMICs where income generated through the informal economy is hard to measure (69) and where healthcare expenditures are poorly tracked (70). Still, they may be useful when granular data is not available or unreliable. Inclusion of such data should be considered in future studies of bacterial infection.

Despite interest in the role of improved WASH in reducing global AMR, we identified only four studies that reported individuals’ colonization/infection with priority bacterial pathogens stratified by their WASH access. This may be a consequence of our search strategy, as WASH access may not typically be considered an SES characteristic and thus relevant studies may not have been identified through our search terms. Alternatively, it could be a consequence of the fact that many recent studies have investigated relationships between AMR and WASH at regional or country-level scales (71) rather than at the individual level.

### Future Directions and Conclusion

This scoping review found sufficient evidence to support future systematic reviews and meta-analyses evaluating the relationship between SES and risks for colonization/infection with community-acquired bacterial pathogens. Nevertheless, we identified several gaps and opportunities for future research. First, we identified few studies from LMICs, where the burden of AMR is highest. Bacterial infections and their antibiotic susceptibility profiles may be poorly characterized in some LMICs due to limited laboratory infrastructure and diagnostic challenges (72). However, the U.S. CDC and other public health organizations are actively working with hospitals and health ministries in LMICs to improve their capacity for timely diagnosis. Expanded laboratory infrastructure in the next several years should provide new opportunities to determine how rates of colonization/infection with priority bacterial pathogens differ within LMIC settings based on the SDOH. This is important to allow intervention efforts focused on hygiene, health practices, and health education to be equally effective across groups. Second, although we report findings by unique, individual SES characteristics, many SES characteristics are collinear by nature. We noted that only a fraction of published studies reported data stratified by SES, as SES is more often controlled for rather than analyzed as an exposure of interest in bacterial colonization/infection studies. Of the studies that did report results stratified by SES, few examined collinearity between reported SES characteristics, making it challenging to assess the most important exposures driving or mediating observed associations. Future studies should report data stratified by SES characteristics or SES deprivation scores to allow for a better understanding of the complex interplay between SES and health, especially in LMICs. As scientists and clinicians, it is imperative that we promptly ascertain the scale of underlying disparities both nationally and globally.

## Data Availability

All data produced in the present work are contained in the manuscript.

## Author Contributions

MLN, CWC, and SD conceptualized the study. MLN, NN, and SD acquired the funding. RM, SD, NN, and CWC performed the literature search. SAB, EA, NN, CWC, S Balaji, LM, SAA, SD, and MLN reviewed titles and abstracts, reviewed full texts, designed the data extraction template, and extracted data. SAB and S Balaji prepared the tables and figures. SAB, RM, and MLN wrote the first draft of the manuscript. All authors reviewed the manuscript.

## Funding

Research reported in this publication was supported by the National Institute of Allergy and Infectious Diseases of the National Institutes of Health under Award Number UM1AI104681. The content is solely the responsibility of the authors and does not necessarily represent the official views of the National Institutes of Health. Funders had no role in study design; in the collection, analysis, or interpretation of data; on in writing the manuscript. CWC was supported by an IDSA Foundation and HIV Medicine Association Grants for Emerging Research/Clinician Mentorship (G.E.R.M.) Program Award. The ARLG Publications Committee reviewed the manuscript prior to submission for publication.

## Notes

### Competing Interest Statement

The authors have declared no competing interest.

## References

1. EClinicalMedicine null. Antimicrobial resistance: a top ten global public health threat. EClinicalMedicine. 2021 Nov;41:101221.

2. Pei S, Blumberg S, Vega JC, Robin T, Zhang Y, Medford RJ, et al. Challenges in Forecasting Antimicrobial Resistance - Volume 29, Number 4—April 2023 - Emerging Infectious Diseases journal - CDC. [cited 2023 Nov 7]; Available from: https://wwwnc.cdc.gov/eid/article/29/4/22-1552_article

3. Global burden of bacterial antimicrobial resistance in 2019: a systematic analysis - The Lancet [Internet]. [cited 2023 Oct 24]. Available from: https://www.thelancet.com/journals/lancet/article/PIIS0140-6736(21)02724-0/fulltext#seccestitle190

4. Social Determinants of Health - Healthy People 2030 | health.gov [Internet]. [cited 2023 Nov 7]. Available from: https://health.gov/healthypeople/priority-areas/social-determinants-health

5. Avendano EE, Blackmon SA, Nirmala N, Chan CW, Morin RA, Balaji S, et al. Race and ethnicity as risk factors for colonization and infection with key bacterial pathogens: a scoping review. Submiss.

6. Covidence systematic review software [Internet]. Melbourne, Australia: Veritas Health Innovation; Available from: www.covidence.org

7. R Core Team. R (4.3.1). A language and environment for statistical computing. R Foundation for Statistical Computing [Internet]. Vienna, Austria; 2023. Available from: https://www.r-project.org/

8. Wickham H. ggplot2: elegant graphics for data analysis. Second edition. Switzerland: Springer; 2016. 260 p. (Use R!).

9. Alsan M, Kammili N, Lakshmi J, Xing A, Khan A, Rani M, et al. Poverty and Community-Acquired Antimicrobial Resistance with Extended-Spectrum β-Lactamase–Producing Organisms, Hyderabad, India. Emerg Infect Dis. 2018 Aug;24(8):1490–6.

10. Como-Sabetti KJ, Harriman KH, Fridkin SK, Jawahir SL, Lynfield R. Risk factors for community-associated *Staphylococcus aureus* infections: results from parallel studies including methicillin-resistant and methicillin-sensitive *S. aureus* compared to uninfected controls. Epidemiol Infect. 2011 Mar;139(3):419–29.

11. Dao TT, Liebenthal D, Tran TK, Ngoc Thi Vu B, Ngoc Thi Nguyen D, Thi Tran HK, et al. Klebsiella pneumoniae Oropharyngeal Carriage in Rural and Urban Vietnam and the Effect of Alcohol Consumption. Kluytmans J, editor. PLoS ONE. 2014 Mar 25;9(3):e91999.

12. GebreSilasie YM, Tullu KD, Yeshanew AG. Resistance pattern and maternal knowledge, attitude and practices of suspected Diarrheagenic Escherichia coli among children under 5 years of age in Addis Ababa, Ethiopia: cross sectional study. Antimicrob Resist Infect Control. 2018 Sep 12;7(1):110.

13. Graham PL, Lin SX, Larson EL. A U.S. Population-Based Survey of *Staphylococcus aureus* Colonization. Ann Intern Med. 2006 Mar 7;144(5):318.

14. Herindrainy P, Randrianirina F, Ratovoson R, Ratsima Hariniana E, Buisson Y, Genel N, et al. Rectal Carriage of Extended-Spectrum Beta-Lactamase-Producing Gram-Negative Bacilli in Community Settings in Madagascar. Paul RE, editor. PLoS ONE. 2011 Jul 29;6(7):e22738.

15. Koch K, Sogaard M, Norgaard M, Thomsen RW, Schonheyder HC, for the Danish Collaborative Bacteremia Network. Socioeconomic Inequalities in Risk of Hospitalization for Community-Acquired Bacteremia: A Danish Population-Based Case-Control Study. Am J Epidemiol. 2014 May 1;179(9):1096–106.

16. Kurowski KM, Marusinec R, Amato HK, Saraiva-Garcia C, Loayza F, Salinas L, et al. Social and Environmental Determinants of Community-Acquired Antimicrobial-Resistant Escherichia coli in Children Living in Semirural Communities of Quito, Ecuador. Am J Trop Med Hyg. 2021 Jul 19;105(3):600–10.

17. Mechal T, Hussen S, Desta M. Bacterial Profile, Antibiotic Susceptibility Pattern and Associated Factors Among Patients Attending Adult OPD at Hawassa University Comprehensive Specialized Hospital, Hawassa, Ethiopia. Infect Drug Resist. 2021 Jan;Volume 14:99–110.

18. Nomamiukor BO, Horner C, Kirby A, Hughes GJ. Living conditions are associated with increased antibiotic resistance in community isolates of Escherichia coli. J Antimicrob Chemother. 2015 Nov;70(11):3154–8.

19. Pathak A, Marothi Y, Iyer RV, Singh B, Sharma M, Eriksson B, et al. Nasal Carriage and Antimicrobial Susceptibility of Staphylococcus aureusin healthy preschool children in Ujjain, India. BMC Pediatr. 2010 Dec;10(1):100.

20. Ravishankar A, Singh S, Rai S, Sharma N, Gupta S, Thawani R. Socio-economic profile of patients with community-acquired skin and soft tissue infections in Delhi. Pathog Glob Health. 2014 Sep;108(6):279–82.

21. See I, Wesson P, Gualandi N, Dumyati G, Harrison LH, Lesher L, et al. Socioeconomic Factors Explain Racial Disparities in Invasive Community-Associated Methicillin-Resistant Staphylococcus aureus Disease Rates. Clin Infect Dis. 2017 Mar 1;64(5):597–604.

22. Beresin GA, Wright JM, Rice GE, Jagai JS. Swine exposure and methicillin-resistant Staphylococcus aureus infection among hospitalized patients with skin and soft tissue infections in Illinois: A ZIP code-level analysis. Environ Res. 2017 Nov;159:46–60.

23. Sardá V, Trick WE, Zhang H, Schwartz DN. Spatial, Ecologic, and Clinical Epidemiology of Community-Onset, Ceftriaxone-Resistant Enterobacteriaceae, Cook County, Illinois, USA. Emerg Infect Dis. 2021 Aug;27(8):2127–34.

24. Fritz SA, Garbutt J, Elward A, Shannon W, Storch GA. Prevalence of and Risk Factors for Community-Acquired Methicillin-Resistant and Methicillin-Sensitive *Staphylococcus aureus* Colonization in Children Seen in a Practice-Based Research Network. Pediatrics. 2008 Jun 1;121(6):1090–8.

25. Ali F, Immergluck LC, Leong T, Waller L, Malhotra K, Jerris RC, et al. A Spatial Analysis of Health Disparities Associated with Antibiotic Resistant Infections in Children Living in Atlanta (2002–2010). EGEMs Gener Evid Methods Improve Patient Outcomes. 2019 Sep 12;7(1):50.

26. Bar-Meir M, Tan TQ. Staphylococcus aureus Skin and Soft Tissue Infections: Can We Anticipate the Culture Result? Clin Pediatr (Phila). 2010 May;49(5):432–8.

27. Berens P, Swaim L, Peterson B. Incidence of Methicillin-Resistant *Staphylococcus aureus* in Postpartum Breast Abscesses. Breastfeed Med. 2010 Jun;5(3):113–5.2

28. Casey JA, Rudolph KE, Robinson SC, Bruxvoort K, Raphael E, Hong V, et al. Sociodemographic Inequalities in Urinary Tract Infection in 2 Large California Health Systems. Open Forum Infect Dis. 2021 Jun 1;8(6):ofab276.

29. Dilks SA, Schlager T, Kopco JA, Lohr JA, Gressard RP, Hendley JO, et al. Frequency and Correlates of Bacteriuria Among Children With Neurogenic Bladder: South Med J. 1993 Dec;86(12):1372–5.

30. Frei CR, Makos BR, Daniels KR, Oramasionwu CU. Emergence of community-acquired methicillin-resistant Staphylococcus aureus skin and soft tissue infections as a common cause of hospitalization in United States children. J Pediatr Surg. 2010 Oct;45(10):1967–74.

31. Immergluck LC, Leong T, Malhotra K, Parker TC, Ali F, Jerris RC, et al. Geographic surveillance of community associated MRSA infections in children using electronic health record data. BMC Infect Dis. 2019 Dec;19(1):170.

32. Len KA, Bergert L, Patel S, Melish M, Kimata C, Erdem G. Community-acquired *Staphylococcus aureus* pneumonia among hospitalized children in Hawaii: MRSA and MSSA Pneumonia in Children. Pediatr Pulmonol. 2010 Sep;45(9):898–905.

33. Mork RL, Hogan PG, Muenks CE, Boyle MG, Thompson RM, Sullivan ML, et al. Longitudinal, strain-specific Staphylococcus aureus introduction and transmission events in households of children with community-associated meticillin-resistant S aureus skin and soft tissue infection: a prospective cohort study. Lancet Infect Dis. 2020 Feb;20(2):188–98.

34. Farr AM, Marx MA, Weiss D, Nash D. Association of neighborhood-level factors with hospitalization for community-associated methicillin-resistant Staphylococcus aureus, New York City, 2006: a multilevel observational study. BMC Infect Dis. 2013 Dec;13(1):84.

35. Goud R, Gupta S, Neogi U, Agarwal D, Naidu K, Chalannavar R, et al. Community prevalence of methicillin and vancomycin resistant Staphylococcus aureus in and around Bangalore, southern India. Rev Soc Bras Med Trop. 2011;44(3):309–12.

36. Mainous AG. Nasal Carriage of Staphylococcus aureus and Methicillin-Resistant S aureus in the United States, 2001-2002. Ann Fam Med. 2006 Mar 1;4(2):132–7.

37. Nerby JM, Gorwitz R, Lesher L, Juni B, Jawahir S, Lynfield R, et al. Risk Factors for Household Transmission of Community-associated Methicillin-resistant Staphylococcus aureus. Pediatr Infect Dis J. 2011 Nov;30(11):927–32.

38. Ray GT, Suaya JA, Baxter R. Incidence, microbiology, and patient characteristics of skin and soft-tissue infections in a U.S. population: a retrospective population-based study. BMC Infect Dis. 2013 Dec;13(1):252.

39. Beltrán MA, García Del Corro HJ, Couso M, Gallo MD, Lettieri A, Barna PV. [Relationship between crowding and community acquired skin and soft tissues infections]. Medicina (Mex). 2017;77(6):465–8.

40. Cohen AL, Shuler C, McAllister S, Fosheim GE, Brown MG, Abercrombie D, et al. Methamphetamine Use and Methicillin-Resistant *Staphylococcus aureus* Skin Infections. Emerg Infect Dis. 2007 Nov;13(11):1707–13.

41. Fritz SA, Hogan PG, Hayek G, Eisenstein KA, Rodriguez M, Krauss M, et al. Staphylococcus aureus Colonization in Children With Community-Associated Staphylococcus aureus Skin Infections and Their Household Contacts. Arch Pediatr Adolesc Med [Internet]. 2012 Jun 1 [cited 2023 Aug 31];166(6). Available from: http://archpedi.jamanetwork.com/article.aspx?doi=10.1001/archpediatrics.2011.900

42. Hobbs MR, Grant CC, Thomas MG, Berry S, Morton SMB, Marks E, et al. Staphylococcus aureus colonisation and its relationship with skin and soft tissue infection in New Zealand children. Eur J Clin Microbiol Infect Dis. 2018 Oct;37(10):2001–10.

43. Hota B. Community-Associated Methicillin-Resistant Staphylococcus aureus Skin and Soft Tissue Infections at a Public Hospital: Do Public Housing and Incarceration Amplify Transmission? Arch Intern Med. 2007 May 28;167(10):1026.

44. Popovich KJ, Smith KY, Khawcharoenporn T, Thurlow CJ, Lough J, Thomas G, et al. Community-Associated Methicillin-Resistant Staphylococcus aureus Colonization in High-Risk Groups of HIV-Infected Patients. Clin Infect Dis. 2012 May 1;54(9):1296–303.

45. Popovich KJ, Hota B, Aroutcheva A, Kurien L, Patel J, Lyles-Banks R, et al. Community-associated methicillin-resistant Staphylococcus aureus colonization burden in HIV-infected patients. Clin Infect Dis Off Publ Infect Dis Soc Am. 2013 Apr;56(8):1067–74.

46. Popovich KJ, Zawitz C, Weinstein RA, Grasso AE, Hotton AL, Hota B. The Intersecting Epidemics of Human Immunodeficiency Virus, Community-Associated Methicillin-Resistant Staphylococcus aureus, and Incarceration. Open Forum Infect Dis. 2015 Dec;2(4):ofv148.

47. Casey JA, Cosgrove SE, Stewart WF, Pollak J, Schwartz BS. A population-based study of the epidemiology and clinical features of methicillin-resistant *Staphylococcus aureus* infection in Pennsylvania, 2001–2010. Epidemiol Infect. 2013 Jun;141(6):1166–79.

48. Chua KYL, Stewardson AJ. Individual and community predictors of urinary ceftriaxone-resistant Escherichia coli isolates, Victoria, Australia. Antimicrob Resist Infect Control. 2019 Feb 12;8:36.

49. McNamara JF, Harris PNA, Chatfield MD, Paterson DL. Acute Myocardial Infarction and Community-acquired *Staphylococcus aureus* Bloodstream Infection: An Observational Cohort Study. Clin Infect Dis. 2021 Nov 2;73(9):e2647–55.

50. Okechukwu AA, Thairu Y. Bacteria urinary tract infection in HIV-infected children and adolescents in Abuja, Nigeria: a cross-sectional study. Afr J Clin Exp Microbiol. 2019 Aug 22;20(4):306.

51. Vogel AM, Borland A, Van Der Werf B, Morales A, Freeman J, Taylor S, et al. Community-acquired invasive *Staphylococcus aureus* : Uncovering disparities and the burden of disease in Auckland children. J Paediatr Child Health. 2020 Feb;56(2):244–51.

52. Wehrhahn MC, Robinson JO, Pearson JC, O’Brien FG, Tan HL, Coombs GW, et al. Clinical and laboratory features of invasive community-onset methicillin-resistant Staphylococcus aureus infection: a prospective case-control study. Eur J Clin Microbiol Infect Dis Off Publ Eur Soc Clin Microbiol. 2010 Aug;29(8):1025–33.

53. Williamson DA, Ritchie SR, Lennon D, Roberts SA, Stewart J, Thomas MG, et al. Increasing Incidence and Sociodemographic Variation in Community-onset Staphylococcus Aureus Skin and Soft Tissue Infections in New Zealand Children. Pediatr Infect Dis J. 2013 Aug;32(8):923–5.

54. Britton PN, Andresen DN. Paediatric community-associated *Staphylococcus aureus* : A retrospective cohort study: Paediatric *Staphylococcus aureus*. J Paediatr Child Health. 2013 Sep;49(9):754–9.

55. Davoodabadi F, Mobasherizadeh S, Mostafavizadeh K, Shojaei H, Havaei S, Koushki A, et al. Nasal colonization in children with community acquired methicillin-resistant Staphylococcus aureus. Adv Biomed Res. 2016;5(1):86.

56. Hoffmann K, Den Heijer CDJ, George A, Apfalter P, Maier M. Prevalence and resistance patterns of commensal S. aureus in community-dwelling GP patients and socio- demographic associations. A cross-sectional study in the framework of the APRES-project in Austria. BMC Infect Dis. 2015 Dec;15(1):213.

57. Hugbo PG, Okonkwo JO. Potential for local and systemic bacterial infection in some occupational groups in Benin City, Nigeria. Am J Infect Control. 1992 Jun;20(3):126–30.

58. Crum-Cianflone NF, Shadyab AH, Weintrob A, Hospenthal DR, Lalani T, Collins G, et al. Association of Methicillin-Resistant Staphylococcus aureus (MRSA) Colonization With High-Risk Sexual Behaviors in Persons Infected With Human Immunodeficiency Virus (HIV). Medicine (Baltimore). 2011 Nov;90(6):379–89.

59. Collignon P, Beggs JJ, Walsh TR, Gandra S, Laxminarayan R. Anthropological and socioeconomic factors contributing to global antimicrobial resistance: a univariate and multivariable analysis. Lancet Planet Health. 2018 Sep 1;2(9):e398–405.

60. Gebeyehu E, Bantie L, Azage M. Inappropriate Use of Antibiotics and Its Associated Factors among Urban and Rural Communities of Bahir Dar City Administration, Northwest Ethiopia. PLoS ONE. 2015 Sep 17;10(9):e0138179.

61. Alividza V, Mariano V, Ahmad R, Charani E, Rawson TM, Holmes AH, et al. Investigating the impact of poverty on colonization and infection with drug-resistant organisms in humans: a systematic review. Infect Dis Poverty. 2018 Aug 17;7(1):76.

62. Dickman SL, Himmelstein DU, Woolhandler S. Inequality and the health-care system in the USA. The Lancet. 2017 Apr 8;389(10077):1431–41.

63. Mainous AG, Everett CJ, Post RE, Diaz VA, Hueston WJ. Availability of Antibiotics for Purchase Without a Prescription on the Internet. Ann Fam Med. 2009 Sep;7(5):431–5.

64. Laytner LA, Olmeda K, Salinas J, Alquicira O, Nash S, Zoorob R, et al. Acculturation and Subjective Norms Impact Non-Prescription Antibiotic Use among Hispanic Patients in the United States. Antibiotics. 2023 Sep 8;12(9):1419.

65. Cutler D, Lleras-Muney A. Education and Health: Evaluating Theories and Evidence [Internet]. Cambridge, MA: National Bureau of Economic Research; 2006 Jul [cited 2023 Oct 24] p. w12352. Report No.: w12352. Available from: http://www.nber.org/papers/w12352.pdf

66. Hatcher SM, Rhodes SM, Stewart JR, Silbergeld E, Pisanic N, Larsen J, et al. The Prevalence of Antibiotic-Resistant Staphylococcus aureus Nasal Carriage among Industrial Hog Operation Workers, Community Residents, and Children Living in Their Households: North Carolina, USA. Environ Health Perspect. 2017 Apr;125(4):560–9.

67. Phillips RL, Liaw W, Crampton P, Exeter DJ, Bazemore A, Vickery KD, et al. How Other Countries Use Deprivation Indices—And Why The United States Desperately Needs One. Health Aff (Millwood). 2016 Nov;35(11):1991–8.

68. Oakes JM, Rossi PH. The measurement of SES in health research: current practice and steps toward a new approach. Soc Sci Med. 2003 Feb 1;56(4):769–84.

69. Deaton A, Zaidi S. Guidelines for Constructing Consumption Aggregates for Welfare Analysis [Internet]. 2002 [cited 2023 Dec 4]. Available from: http://hdl.handle.net/10986/14101

70. Hendriksen RS, Munk P, Njage P, van Bunnik B, McNally L, Lukjancenko O, et al. Global monitoring of antimicrobial resistance based on metagenomics analyses of urban sewage. Nat Commun. 2019 Mar 8;10(1):1124.

71. Iskandar K, Molinier L, Hallit S, Sartelli M, Hardcastle TC, Haque M, et al. Surveillance of antimicrobial resistance in low- and middle-income countries: a scattered picture. Antimicrob Resist Infect Control. 2021 Mar 31;10:63.

72. CDC Global Health Strategy | Global Health | CDC [Internet]. 2023 [cited 2023 Dec 5]. Available from: https://www.cdc.gov/globalhealth/strategy/default.htm

