## Supplementary Materials for "Socioeconomic status and the risk for colonization or infection with priority bacterial pathogens: a global evidence map"

**Table S1.** Medline search strategy.

**Table S2.** PRISMA-ScR

**Table S3.** Publications that met inclusion criteria.

**Table S1.** Medline search strategy.

| 1 | Community-Acquired Infections/ |
| --- | --- |
| 2 | community acquired.mp. |
| 3 | community associated.mp. |
| 4 | community onset.mp. |
| 5 | Outpatients/ |
| 6 | outpatient$.mp. |
| 7 | Ambulatory Care/ |
| 8 | ambulatory.mp. |
| 9 | 1 or 2 or 3 or 4 or 5 or 6 or 7 or 8 |
| 10 | exp Socioeconomic Factors/ |
| 11 | socioeconomic$.mp. |
| 12 | socio-economic$.mp. |
| 13 | (standard$ adj2 living).mp. |
| 14 | social inequit$.mp. |
| 15 | social inequalit$.mp. |
| 16 | Health Status Disparities/ |
| 17 | Healthcare Disparities/ |
| 18 | (Health$ adj3 Disparit$).mp. |
| 19 | exp continental population groups/ or exp ethnic groups/ |
| 20 | (race$ or racial).mp. |
| 21 | ethnic$.mp. |
| 22 | 10 or 11 or 12 or 13 or 14 or 15 or 16 or 17 or 18 or 19 or 20 or 21 |
| 23 | Enterococcus faecium/ |
| 24 | exp Staphylococcus aureus/ |
| 25 | exp Staphylococcal Infections/ |
| 26 | Klebsiella pneumoniae/ |
| 27 | Acinetobacter baumannii/ |
| 28 | Pseudomonas aeruginosa/ |
| 29 | exp Enterobacter/ |
| 30 | exp Escherichia coli/ |
| 31 | exp Citrobacter/ |
| 32 | Enterobacteriaceae/ |
| 33 | Enterococcus faecium.mp. |
| 34 | Staphylococcus aureus.mp. |
| 35 | Klebsiella pneumoniae.mp. |
| 36 | Acinetobacter baumannii.mp. |
| 37 | Pseudomonas aeruginosa.mp. |
| 38 | Enterobacter.mp. |
| 39 | Escherichia coli.mp. |
| 40 | E coli.mp. |
| 41 | Citrobacter.mp. |
| 42 | gram-negative bacterial infections/ |
| 43 | exp enterobacteriaceae infections/ |
| 44 | exp moraxellaceae infections/ |
| 45 | Gram-Negative Bacteria/ |
| 46 | gram negative.mp. |
| 47 | Enterobacteriaceae.mp. |
| 48 | ESKAPE.mp. |
| 49 | 23 or 24 or 25 or 26 or 27 or 28 or 29 or 30 or 31 or 32 or 33 or 34 or 35   or 36 or 37 or 38 or 39 or 40 or 41 or 42 or 43 or 44 or 45 or 46 or 47   or 48 |
| 50 | 9 and 22 and 49 |

**Table S2.** PRISMA-ScR Checklist

| **SECTION** | **ITEM** | **PRISMA-ScR CHECKLIST ITEM** | **REPORTED ON PAGE #** |
| --- | --- | --- | --- |
| **TITLE** | | | |
| Title | 1 | Identify the report as a scoping review. | 1 |
| **ABSTRACT** | | | |
| Structured summary | 2 | Provide a structured summary that includes (as applicable): background, objectives, eligibility criteria, sources of evidence, charting methods, results, and conclusions that relate to the review questions and objectives. | 2 |
| **INTRODUCTION** | | | |
| Rationale | 3 | Describe the rationale for the review in the context of what is already known. Explain why the review questions/objectives lend themselves to a scoping review approach. | 4 |
| Objectives | 4 | Provide an explicit statement of the questions and objectives being addressed with reference to their key elements (e.g., population or participants, concepts, and context) or other relevant key elements used to conceptualize the review questions and/or objectives. | 4 |
| **METHODS** | | | |
| Protocol and registration | 5 | Indicate whether a review protocol exists; state if and where it can be accessed (e.g., a Web address); and if available, provide registration information, including the registration number. | 5 |
| Eligibility criteria | 6 | Specify characteristics of the sources of evidence used as eligibility criteria (e.g., years considered, language, and publication status), and provide a rationale. | 5 |
| Information sources* | 7 | Describe all information sources in the search (e.g., databases with dates of coverage and contact with authors to identify additional sources), as well as the date the most recent search was executed. | 5 |
| Search | 8 | Present the full electronic search strategy for at least 1 database, including any limits used, such that it could be repeated. | 5 |
| Selection of sources of evidence† | 9 | State the process for selecting sources of evidence (i.e., screening and eligibility) included in the scoping review. | 5 |
| Data charting process‡ | 10 | Describe the methods of charting data from the included sources of evidence (e.g., calibrated forms or forms that have been tested by the team before their use, and whether data charting was done independently or in duplicate) and any processes for obtaining and confirming data from investigators. | 5-6 |
| Data items | 11 | List and define all variables for which data were sought and any assumptions and simplifications made. | 6 |
| Critical appraisal of individual sources of evidence§ | 12 | If done, provide a rationale for conducting a critical appraisal of included sources of evidence; describe the methods used and how this information was used in any data synthesis (if appropriate). | - |
| Synthesis of results | 13 | Describe the methods of handling and summarizing the data that were charted. | 6 |
| **RESULTS** | | | |
| Selection of sources of evidence | 14 | Give numbers of sources of evidence screened, assessed for eligibility, and included in the review, with reasons for exclusions at each stage, ideally using a flow diagram. | 6 |
| Characteristics of sources of evidence | 15 | For each source of evidence, present characteristics for which data were charted and provide the citations. | 6 |
| Critical appraisal within sources of evidence | 16 | If done, present data on critical appraisal of included sources of evidence (see item 12). | - |
| Results of individual sources of evidence | 17 | For each included source of evidence, present the relevant data that were charted that relate to the review questions and objectives. | 6 |
| Synthesis of results | 18 | Summarize and/or present the charting results as they relate to the review questions and objectives. | 6-10 |
| **DISCUSSION** | | | |
| Summary of evidence | 19 | Summarize the main results (including an overview of concepts, themes, and types of evidence available), link to the review questions and objectives, and consider the relevance to key groups. | 10-12 |
| Limitations | 20 | Discuss the limitations of the scoping review process. | 13 |
| Conclusions | 21 | Provide a general interpretation of the results with respect to the review questions and objectives, as well as potential implications and/or next steps. | 13 |
| **FUNDING** | | | |
| Funding | 22 | Describe sources of funding for the included sources of evidence, as well as sources of funding for the scoping review. Describe the role of the funders of the scoping review. | 13 |

JBI = Joanna Briggs Institute; PRISMA-ScR = Preferred Reporting Items for Systematic reviews and Meta-Analyses extension for Scoping Reviews.

* Where *sources of evidence* (see second footnote) are compiled from, such as bibliographic databases, social media platforms, and Web sites.

† A more inclusive/heterogeneous term used to account for the different types of evidence or data sources (e.g., quantitative and/or qualitative research, expert opinion, and policy documents) that may be eligible in a scoping review as opposed to only studies. This is not to be confused with *information sources* (see first footnote).

‡ The frameworks by Arksey and O’Malley (6) and Levac and colleagues (7) and the JBI guidance (4, 5) refer to the process of data extraction in a scoping review as data charting*.*

§ The process of systematically examining research evidence to assess its validity, results, and relevance before using it to inform a decision. This term is used for items 12 and 19 instead of "risk of bias" (which is more applicable to systematic reviews of interventions) to include and acknowledge the various sources of evidence that may be used in a scoping review (e.g., quantitative and/or qualitative research, expert opinion, and policy document).

**Table S3.** Publications that met inclusion criteria.

| Author, Year,  PMID | Country | Study Duration | Funding Source | SES Indicator(s) | Age Group | Infection/Colonization | Recruitment  Method/Data Source^a^ | Bacteria Strain | Outcome | Citation^b^ |
| --- | --- | --- | --- | --- | --- | --- | --- | --- | --- | --- |
| Ali 2019  31565665 | United states | 8 years | Academia, Federal, Other | Income, Healthcare access, Residential crowding | Pediatric | Infection | Medical records | Staphylococcus aureus | SSTI; uTBI | 24 |
| Alsan 2018  30014842 | India | 1 year | Academia | Education, Income,  WASH | Adult | Colonization | Outpatient | Staphylococcus aureus; Klebsiella pneumoniae;  Enterobacter species; Escherichia coli | Bacteriuria (>10^5 CFU/mL bacteria in urine) | 17 |
| Bar-Meir 2010  20118096 | United states | 18 months | None | Healthcare access | Pediatric | Infection | Inpatient | Staphylococcus aureus | SSTI | 25 |
| Beltrán 2017  29223936 | Argentina | 37 days | NR | Residential crowding, SES deprivation score | Adult | Infection | Inpatient,  Outpatient | Staphylococcus aureus | SSTI | 42 |
| Berens 2010  20113200 | United states | 7 years | NR | Healthcare access | Adult | Infection | Medical records | Staphylococcus aureus | Breast abscess | 26 |
| Beresin 2017  28772149 | United  States | 3 years 7 months | Federal, Non profit | Income, Healthcare access, SES deprivation score,  Urbanicity | Adult, Pediatric | Infection | Medical records | Staphylococcus aureus | SSTI | 27 |
| Britton 2013  23721234 | Australia | 1 year | None | Urbanicity | Pediatric | Infection | Medical records | Staphylococcus aureus | SSTI | 56 |
| Casey 2013  22929058 | United states | 9 years | Academia, Federal | SES deprivation score, Urbanicity | Adult, pediatric,  Senior | Infection | Medical records | Staphylococcus aureus | SSTI | 55 |
| Casey 2021  34189179 | United states | 3 years | Federal | Healthcare access, SES deprivation score, Urbanicity | Adult | Infection | Medical records | Escherichia coli | UTI | 28 |
| Chua 2019  30805183 | Australia | 1 month | NR | SES deprivation score, Urbanicity | Adult, pediatric,  Senior | Infection | Inpatient,  Outpatient, Communitybased | Escherichia coli | UTI | 51 |
| Cohen 2007  18217555 | United states | 56 days | NR | Residential crowding | Adult, pediatric,  Senior | Infection | Outpatient | Staphylococcus aureus | SSTI | 44 |
| Como-Sabetti  2011 20513251 | United states | 3 years | Federal | Income, Education, Residential crowding, | Adult,Pediatric,  Senior | Infection | Medical records | Staphylococcus aureus | SSTI; Non-SSTI/non-invasive (ear, eye, urine), and invasive (blood, joint, bone) | 20 |
| Crum-Cianflone  2011 24667800 | United  States | 3 years | Federal | WASH | Adult | Colonization | Outpatient | Klebsiella pneumoniae | Colonized in oropharyngeal tract  (either nose, throat, or both) | 60 |

| Dao 2014  24667800 | Vietnam | NR | Non- profit | | Income, Education,  Urbanicity | | Adult, pediatric | | Colonization | | Households | | Staphylococcus aureus | Swab from nares, throat, bilateral axilla areas, bilateral groin areas, and perirectal area | 11 | |
| --- | --- | --- | --- | --- | --- | --- | --- | --- | --- | --- | --- | --- | --- | --- | --- | --- |
| Davoodabadi  2016 27274501 | Iran | 13 months | None | | Urbanicity | | Pediatric | | Colonization | | Outpatient | | Staphylococcus aureus | Nare swab | 58 | |
| Dilks 1993  8272914 | United  States | 2 years | NR | | Healthcare access | | Pediatric | | Colonization | | Outpatient | | Klebsiella pneumoniae; Escherichia coli | Bacteriuria | 29 | |
| Farr 2013  23406159 | United states | 1 year | None | | Income | | Adult,Pediatric,  Senior | | Infection | | Medical records | | Staphylococcus aureus | MRSA hospitalization | 35 | |
| Frei 2010  20920714 | United  States | 11 years | Academia | | Healthcare access | | Pediatrics | | Infection | | Inpatient | | Staphylococcus aureus | SSTI | 30 | |
| Fritz 2008  18519477 | United states | 8 months | Federal | | Healthcare access, Residential crowding | | Pediatric | | Colonization | | Outpatient | | Staphylococcus aureus | Nasal swabs | 23 | |
| Fritz 2012  22665030 | United states | 20 months | Academia, Federal, Non-profit | | Residential crowding | | Adult,Pediatric,  Senior | | Colonization | | Households | | Staphylococcus aureus | Swab from anterior nares, axillae, inguinal folds | 45 | |
| GebreSilasie  2018 30214719 | Ethiopia | 5 months | Academia | | Education, Income,  WASH | | Pediatric | | Infection | | Outpatient | | Escherichia coli | Diarrhea; Suspected Diarrheagenic  E. coli | 15 | |
| Goud 2011  21901873 | India | 4 years 8 months | NR | | Income | | Adult,Pediatric | | Colonization | | Inpatient,  Communitybased | | Staphylococcus aureus | Staphylococci colonization of anterior nares, forearm, dorsum and palm of the hands | 34 | |
| Graham 2006  16520472 | United states | 1 year | Federal | | Education,  Healthcare access | | Adult,Pediatric,  Senior | | Colonization | | Community based | | Staphylococcus aureus | Nasal colonization | 21 | |
| Herindrainy  2011 21829498 | Madagascar | 5 months | Non- profit | | Education, Residential crowding | | Adult | | Infection | | Outpatient | | Klebsiella pneumoniae; Enterobacter species;  Escherichia coli | Intestinal carriage of ESBL-PE | 10 | |
| Hobbs 2018  30066280 | New  Zealand | 4.5 years | Academia, industry, Federal, Other-  Auckland Medical Research  Foundation | | Residential crowding, SES deprivation score,  Urbanicity | | Pediatric | | Colonization | | Community based | | Staphylococcus aureus | SSTI: S. aureus colonization (swab from nasal, oropharynx, skin) | 41 | |
| Hoffmann 2015  25981559 | Austria | 9 months | Federal | | Urbanicity | | Adult,Pediatric,  Senior | | Colonization | | Outpatient | | Staphylococcus aureus | Nasal Swabs | 57 | |
| Hota 2007  17533205 | United states | 5 years and  8 months | None | | Residential crowding | | Adult | | Infection | | Medical records | | Staphylococcus aureus | SSTI | 43 | |
| Hugbo 1992  1636931 | Nigeria | 6 months | | NR | | Urbanicity | | Adult,Pediatric | | Colonization | Outpatient,  Community based | Staphylococcus aureus; Escherichia coli | | Swab taken from back of palm, cheek, anterior nares, ear entrance,  Colonization with pathogens | | 59 |
| Immergluck  2019 30777016 | United  States | 9 years | | Academia, Federal, Non-profit | | Income, Healthcare access, Residential crowding | | Pediatric | | Infection | Medical records | Staphylococcus aureus | | SSTI | | 31 |
| Koch 2014  24682527 | Denmark | 8 years | | Academia | | Education, Income | | Adult | | Infection | Medical records | Enterococcus faecium; Staphylococcus aureus;  Pseudomonas aeruginosa; Enterobacter species; Escherichia coli | | Bacteremia | | 9 |
| Kurowski 2021  34280150 | Ecuador | 10 months | | Federal | | Education, Income, Residential crowding, WASH | | Pediatric | | Colonization | Households | Escherichia coli | | Fecal samples | | 13 |
| Len 2010  20632405 | United  States | 12 years | | None | | Healthcare access | | Pediatric | | Infection | Inpatient | Staphylococcus aureus | | SSTI | | 32 |
| Mainous 2006  16569716 | United  States | 2 years | | Federal, Non profit | | Income | | Adult,Pediatric,  Senior | | Colonization | Community based | Staphylococcus aureus | | Nasal carriage of Staphylococcus aureus | | 36 |
| McNamara  2021 32797225 | Australia | 11 years | | Federal | | SES deprivation score | | Adult, Senior | | Infection | Medical records | Staphylococcus aureus | | Bloodstream Infection (BSI) | | 49 |
| Mechal 2021  33469325 | Ethiopia | 5 months | | None | | Education | | Adult | | Infection | Outpatient | Staphylococcus aureus; Klebsiella pneumoniae; Pseudomonas aeruginosa; Enterobacter species; Escherichia coli | | UTI | | 14 |
| Mork 2020  31784369 | United  States | 3 years | | Federal | | Healthcare access | | Adult | | Colonization | Community based | Staphylococcus aureus | | S. aureus transmission or introduction | | 33 |
| Nerby 2011  21617572 | United  States | 1 year and 9 months | | Federal | | Income | | Pediatric | | Colonization | Medical records | Staphylococcus aureus | | Nasal swabs | | 37 |
| Nomamiukor  2015 26260128 | United  Kingdom | 3 years | | Federal | | Education, Income,  SES deprivation score, Urbanicity | | not given | | Infection | Medical records | Escherichia coli | | UTI | | 12 |
| Okechukwu  2019 No PMID | Nigeria | 6 months | | NR | | SES deprivation score | | Pediatric | | Infection | Outpatient | Staphylococcus aureus; Klebsiella pneumoniae;  Pseudomonas aeruginosa; Escherichia coli | | UTI | | 54 |
| Pathak 2010  21190550 | India | 15 months | | Federal | | Education | | Pediatric | | Colonization | Outpatient | Staphylococcus aureus | | SA nasal carriage | | 18 |
| Popovich 2012  22354926 | United  States | 6 months | | Federal | | Residential crowding, SES deprivation score | | Adult | | Colonization | Outpatient | Staphylococcus aureus | | MRSA nasal colonization | | 46 |

| Popovich 2013  23325428 | United states | 13 months | Federal | Residential crowding | Adult | Colonization | Medical records | Staphylococcus aureus | Swab specimens were obtained from the nares, throat, bilateral axillae, bilateral inguinal regions, peri-rectal area, and a chronic  wound if present | 48 |
| --- | --- | --- | --- | --- | --- | --- | --- | --- | --- | --- |
| Popovich 2015  26543878 | United states | 6 years | Federal | Residential crowding | Adult | Infection | Medical records | Staphylococcus aureus | Clinical cultures from skin and soft tissue sites | 47 |
| Ravishankar  2014 25292293 | India | 7 months | Short Term Studentship under the  Indian Council of Medical Research  (ICMR) | Education, Income, SES deprivation score | Adult,Pediatric | Infection | Outpatient | Staphylococcus aureus; Acinetobacter baumannii; Pseudomonas aeruginosa; Enterobacter species; Escherichia coli | SSTI | 16 |
| Ray 2013  23721377 | United states | 3 years | Industry | Income | Adult,Pediatric,  Senior | Infection | Medical records | Staphylococcus aureus | SSTI | 38 |
| Sardá 2021  34287121 | United  States | 3 years | NR | Healthcare access,  Income,  Residential crowding | Adult, Senior | Infection | Medical records | Klebsiella pneumoniae; Enterobacter species;  Escherichia coli;  Enterobacteriaceae/Enterobacterales | UTI; SSTI; Blood | 22 |
| See 2017  28362911 | United  States | 3 years | Federal | Education,  Healthcare access,  Income,  Residential crowding, Urbanicity | Adult | Infection | Medical records | Staphylococcus aureus | Sterile body site (e.g. blood, cerebrospinal fluiud, internal body fluid) | 19 |
| Vogel 2020  31355978 | New  Zealand | 5 years | Academia, Other-Academia: University of Auckland summer studentship to A Morales (one author), District Health Board data analysts provided ICD data. So, pretty much unfunded. | SES deprivation score | Pediatric | Infection | Medical records | Staphylococcus aureus | Invasive S. aureus infection (iSA) | 52 |
| Wehrhahn  2010 20549534 | Australia | 2 years | Non-profit | SES deprivation score | Adult, Pediatric | Infection | Medical records | Staphylococcus aureus | CAP; SSTI; Musculoskeletal infection (Osteomyelitis, Septic arthritis, Bursitis, Tenosynovitis, Epidural abscess), Bloodstream infection (BSI), Infective endocarditisis (IE) | 50 |
| Williamson  2013 23518822 | New  Zealand | 4 years | Internal funding | SES deprivation score | Pediatric | Infection | Medical records | Staphylococcus aureus | SSTI | 53 |

^a^Outpatient settings refer to physical places (I.e., doctors' offices, clinics, etc.) where patients were recruited. Medical records refer to recruitment through medical record or actual data collection was done using medical records.

^b^Number refers to order in references section
